# Astrocytes in neuropathic pain: Mechanistic and global insights

**DOI:** 10.64898/2026.01.23.26344689

**Authors:** Li Xue, Haixia Fan, Hong Yuan, Jiao Yang, Qiurui Yuan

**Author notes:** **(HF)**.

## Abstract

**Background:** Neuropathic pain (NP) is a debilitating chronic pain condition caused by injury or disease of the somatosensory nervous system. Accumulating evidence indicates that astrocytes play a central role in neuroinflammatory regulation and synaptic remodeling, thereby critically influencing the initiation and persistence of neuropathic pain. However, a comprehensive overview of research trends and knowledge structures in this field is still lacking.

**Methods:** The analysis was conducted based on publications retrieved from the Web of Science Core Collection and Scopus, covering the period from 2000 to 2025. Studies focusing on astrocytes and neuropathic pain were systematically identified. Visualization and network analyses were performed using CiteSpace, VOSviewer, and the R package bibliometrix. Collaboration networks, co-citation patterns, keyword co-occurrence, and thematic evolution were analyzed to delineate research hotspots, developmental trajectories, and scholarly contributions across countries, institutions, authors, and journals.

**Results:** 1,828 publications were included, showing a 15% average annual growth in output, which accelerated post-2010. The USA and China led in research and international collaboration, with studies concentrated in North American and East Asian institutions. Author productivity was uneven, with key researchers (Ji RR, Zhang Y, Watkins LR) contributing heavily to publications and citations. *Pain* and *Molecular Pain* were the core journals. Key themes included spinal astrocytic mechanisms, glial activation, and therapeutic modulation, with the focus evolving from injury models/markers to astrocytic activation and targeted pathways.

**Conclusion:** Our analysis shows a substantial growth in astrocyte-related NP research the past 25 years, underscoring astrocytes’ key role in chronic pain pathophysiology. Current trends underscore the integration of mechanistic insights with translational relevance, thereby informing future therapeutic and mechanistic advancements in NP.

## 1. Introduction

Neuropathic pain (NP) is a complex, chronic pain condition that arises from damage to the nervous system, often resulting from injury, disease, or infection[1]. Epidemiological studies indicate that NP affects approximately 7-10% of the general population, with a higher prevalence observed in individuals with conditions such as diabetes, multiple sclerosis, and postherpetic neuralgia. This type of pain can significantly impact quality of life, leading to increased healthcare utilization and associated economic burdens[2].

Astrocytes are increasingly recognized for their pivotal role in the pathophysiology of NP, influencing neuroinflammation, synaptic remodeling, and central sensitization[3]. Following nerve injury, activated astrocytes contribute to neuroinflammatory processes, leading to the production of pro-inflammatory factors that exacerbate pain signaling[4]. Their morphological and transcriptional changes, particularly in the dorsal horn, are associated with the amplification of NP signaling[5]. Additionally, astrocytes interact with microglia, further modulating neuroinflammation and central sensitization, which are key components in the development and maintenance of NP [6]. Targeting astrocytic pathways may offer therapeutic potential for alleviating NP by restoring balance in neuroinflammatory responses and pain signaling mechanisms[7]. Overall, astrocytes are emerging as increasingly critical players in the pathogenesis of NP.

Despite the growing body of literature on astrocytes in NP, a comprehensive overview of research trends and knowledge structures remains limited. Bibliometric analyses reveal an increasing interest in astrocytes across various neurological conditions; however, a focused analysis on NP is lacking [8]. Scientific mapping techniques have been employed in other fields to elucidate complex interactions, indicating a potential method to enhance the understanding of astrocytes in NP[9].

This study employs bibliometric analysis to systematically map the scientific landscape surrounding NP and astrocytes, revealing key research trends and emerging topics in the field. By identifying gaps and opportunities in current investigations, the findings will guide future research efforts aimed at developing targeted therapies that address astrocytic dysfunction in NP.

## 2. Methods

### 2.1 Search strategy

Fig 1 illustrates the process of data retrieval and the criteria used for exclusion. Bibliometric data were collected by applying predefined search terms to the WoSCC and Scopus databases on November 2, 2025. An initial screening was then conducted based on the titles, abstracts, and keywords of the retrieved publications. The selection of search terms was based on Medical Subject Headings (MeSH) for term retrieval. The search strategies used for literature retrieval from the Web of Science Core Collection (WoSCC) and Scopus databases are detailed in S4 Table. By integrating multiple relevant keywords for the initial search, 604 records were retrieved from the WoSCC database, while 1,837 records were obtained from Scopus.

**Fig 1:**
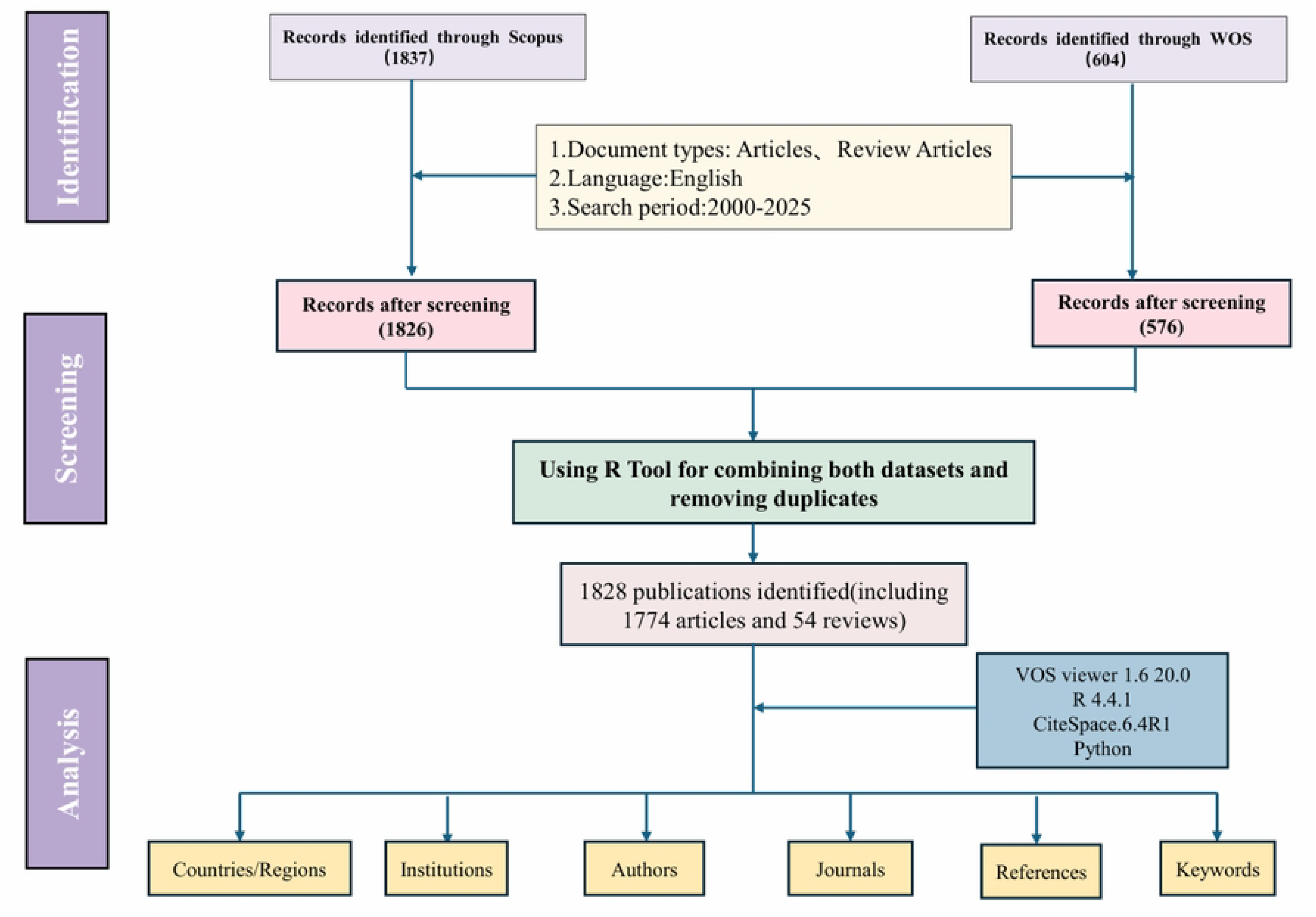
The flowchart for literature search, selection, and analysis.

After data screening, various non-target document types were removed, encompassing proceedings papers, corrections, early access articles, news items, book chapters, retractions, reprints, biographical items, book reviews, meeting abstracts, editorial materials, and letters. The remaining dataset was limited to English-language articles and reviews published between 2000 and 2025.After applying these criteria, 576 records from (WoSCC) and 1,826 records from Scopus were retained (Fig 1).

Subsequently, using the bibliometrix package in R, the two datasets were combined, and duplicate records were subsequently screened based on matching titles, author lists, publication years, and DOIs. When DOIs were unavailable, duplicates were identified using a combination of article titles and first authors. All records flagged as potential duplicates through automated procedures were further examined manually to confirm their validity. This process ensured that publications indexed in both WoSCC and Scopus were counted only once. After this quality control step, a total of 1,828 unique studies were included for subsequent analyses, comprising 1,774 research articles and 54 review papers (Fig 1). These procedures were undertaken to minimize duplication bias and enhance the reliability of the dataset. Two reviewers independently conducted the screening of titles and abstracts, followed by full-text evaluations according to predefined eligibility criteria. Discrepancies were resolved through consensus discussions. After the removal of duplicate records, 1,828 unique studies remained for subsequent analysis.

### 2.2 Bibliometric and scientometric analysis

Bibliometric and scientometric investigations were conducted using multiple complementary software tools to improve the reliability of the analysis. CiteSpace (version 6.4R1, 64-bit Advanced Edition) was utilized to construct visual knowledge maps illustrating authors, institutions, and keywords, employing a one-year time slicing strategy applied across the period from 2000 to 2025[10]. In the analyses of authors and institutions, the top 25 most significant nodes in each time slice were retained. For keyword analysis, pathfinder pruning and network merging techniques were adopted to reduce network complexity and highlight the principal thematic structures. VOSviewer (version 1.6.20) was employed with the full counting method to visualize collaborative networks, including co-authorship patterns and inter-institutional collaborations[11]. Additionally, the R package bibliometrix (https://www.bibliometrix.org)[12] was applied for historiographic analysis and for calculating key bibliometric indicators, such as the g-index[13], h-index[14], number of citations (NC), and number of publications, thereby assessing research productivity and scholarly impact. Microsoft Excel 2021 was employed for initial data sorting and preprocessing.

## 3. Results

### 3.1 Annual publications trends

The literature published between 2000 and 2025 revealed a marked upward trend in both publication output and citation frequency in the research field of astrocytes and NP. A total of 1,828 records were retrieved from the Web of Science Core Collection(WoSCC) and Scopus databases, exhibiting an annual growth rate of 15.24%, which indicates a continuously expanding body of research (S1 Table). As illustrated in Fig 2A, both annual publications and citations derived from the WoSCC saw steady growth in the early years, but there was a noticeable increase, with a sharp rise occurring after 2005.Fig 2B displays the trend from the Scopus database, which aligns with the trends shown in Fig 2A. Fig 2C illustrates the cumulative publication output over time, revealing a pronounced increasing trend, particularly over the past two decades. To quantitatively assess this expansion, a Price’s Law–based growth curve fitting is shown in Fig 2D. The fitted exponential model, expressed as y = 10.114e^0.2289x^, achieves a high coefficient of determination (R² = 0.9209), indicating strong concordance between the model and empirical data. These results substantiate that the publication growth in this field follows an exponential pattern consistent with Price’s Law, reflecting not only a substantial increase in research output but also the dynamic and rapidly advancing nature of the discipline.

**Fig 2:**
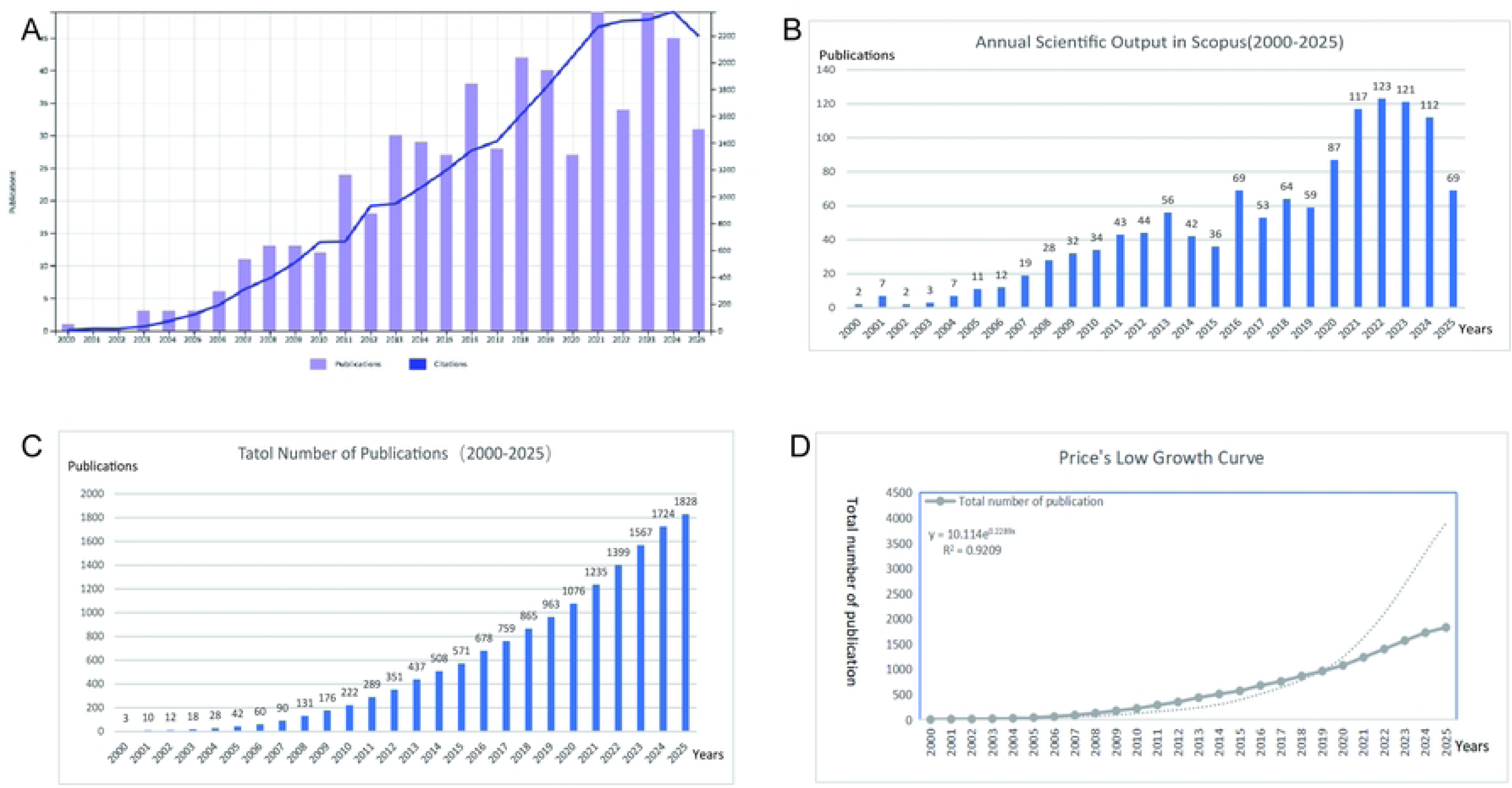
Publication trends in neuropathic pain research from 2000 to 2025. (A) Annual trends in the number of publications (bars) and citations (line) retrieved from the WoSCC database. (B) Annual trends in publications (bars) retrieved from the Scopus database. (C) Cumulative number of publications over time, illustrating the overall growth of the field. (D) Price’s Law curve fitting analysis of cumulative publications, demonstrating the exponential growth pattern of astrocyte-neuropathic pain research.

### 3.2 Distributions of countries/regions

Fig 3A illustrates the global geographical collaboration network, where darker blue shading represents more intensive research ties. The USA and China appear as core nodes, with prominent intercontinental collaboration links, consistent with their leading status in publication output (S2 Table), which shows that China contributed 239 publications (13.1%) and the USA contributed 94 publications (5.1%) in terms of the corresponding authors’ countries of affiliation. In contrast, regions like Africa and South America show sparse connections, reflecting a geographical imbalance in global research partnerships (S2 Table). Fig 3B further visualizes collaborative relationships: the USA. and China form two central clusters, connected to multiple countries (e.g., the UK, Germany, Australia), confirming their role as hubs of international collaboration. Fig 3C quantifies the temporal evolution of research output across key nations: the USA (purple line) maintained a steady upward trajectory in annual publications from the early 2000s onward, while China (red line) exhibited a negligible initial output but a sharp surge post-2010 surge, consistent with expanded funding and infrastructure investment in the field. Other nations (e.g., the ITALY, JAPAN, KOREA, represented by light yellow, blue, and green lines) showed moderate, sustained growth. Remaining secondary to the U.S. and China in annual output by the 2020s, this trend aligns with the cumulative growth pattern observed in S2 Table. Fig 3D (Country Collaboration and Output Distribution Bar Chart) quantifies the research contributions and collaboration engagement of individual countries in the field: The results demonstrated that the USA exhibited significantly higher collaboration intensity than other countries, with the longest red bar among all nations; KOREA and China followed closely in collaboration intensity, and the three countries also showed prominent teal bars for research output — a feature that fully reflects their dual dominance in both research output and international collaboration. In contrast, countries such as Japan and ITALY demonstrate moderate levels of both collaboration intensity and output scale, while nations including the Czech Republic and Finland make relatively limited contributions.

**Fig 3:**
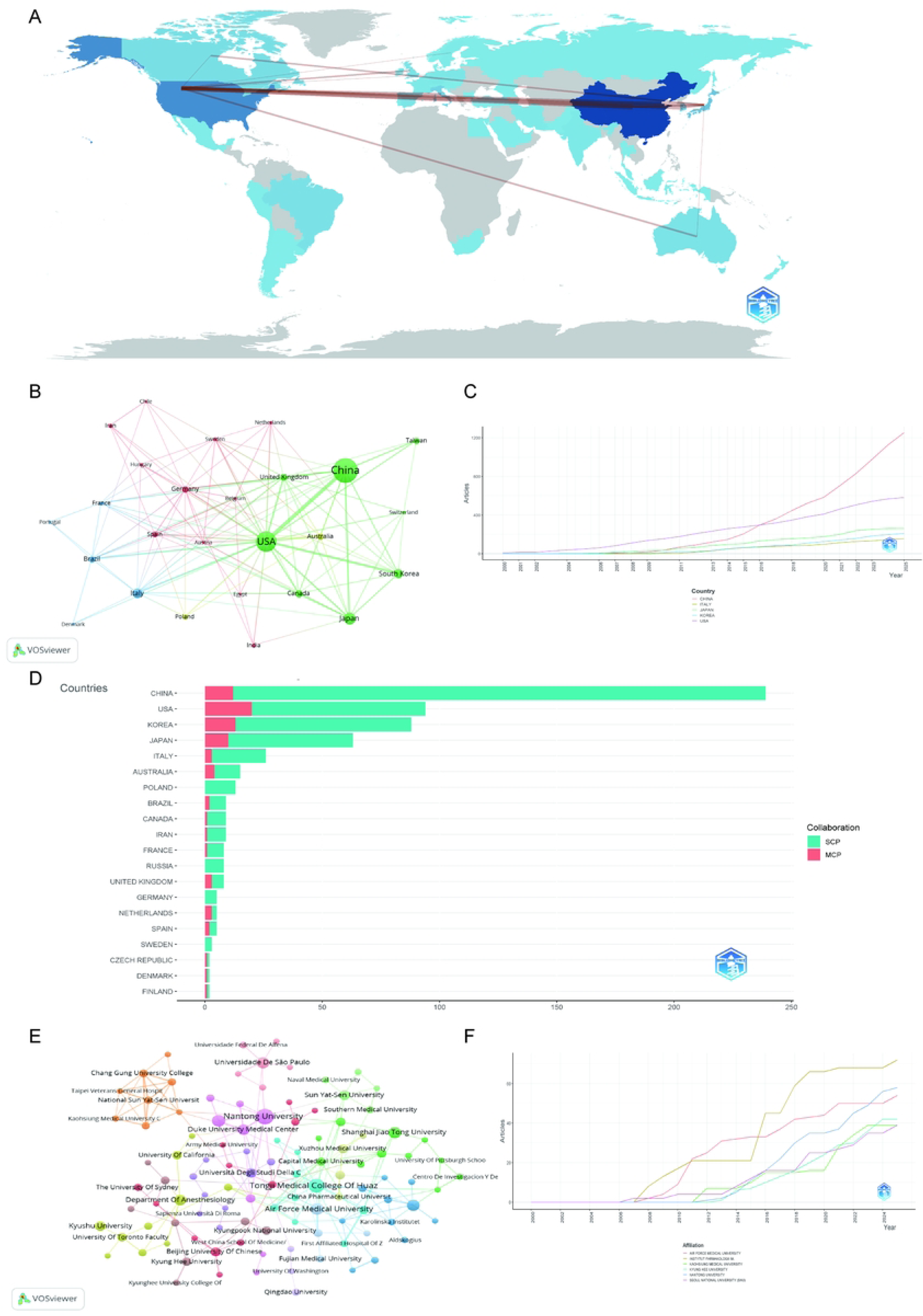
Visualization and analysis of country/region and institutional involvement in neuropathic pain research. (A) Country/Region Collaboration Map. Darker shades of blue indicate higher collaboration rates, and the width of the links represents the strength of collaboration between two countries. (B) Country Clustering Analysis. Each node represents a country or region, with node size proportional to publication output. Edge thickness indicates the strength of co-authorship links between countries, while node colors denote collaboration clusters. (C) Top 5 Countries/Regions by Publications. Lines illustrate the cumulative number of publications from 2000 to 2025. The x-axis represents the year, and the y-axis represents the cumulative document count. (D) Leading countries by the number of studies and collaboration type. Teal segments represent single-country publications (SCP), while red segments represent multiple-country publications (MCP). (E) Institutional collaboration network. Node size corresponds to the extent of collaboration activity, and distinct colors denote clusters, generally reflecting regional or institutional groupings. Line thickness indicates the intensity of cooperation. (F) Institutional publication trends. The x-axis represents years, and the y-axis represents publication counts.

### 3.3 Characteristics of the institutional collaboration network

Fig 3E visualizes the collaborative relationships among institutions in the field, where node size corresponds to research output, and edge connections represent collaborative ties. The network exhibits a multi-cluster structure with geographically and thematically linked institutional groups: A prominent cluster centered on Chinese institutions (e.g., Nantong University, Sun Yat-Sen University, Shanghai Jiao Tong University) forms a dense collaborative sub-network, reflecting intensive domestic partnerships in the field. North American institutions (e.g., University of California, Duke University Medical Center, University of Washington) constitute another key cluster, connected both internally and to select international partners. Scattered nodes (e.g., University of São Paulo [Brazil], Karolinska Institutet [Sweden], University of Sydney [Australia]) represent institutions with relatively independent collaboration patterns, linking to core clusters via limited cross-regional ties. Notably, institutions with larger nodes (e.g., University of California, Sun Yat-Sen University) act as “bridging nodes,” connecting multiple sub-clusters and facilitating knowledge exchange across geographic regions. Fig 3F illustrates the cumulative publication output of key institutions from 2000 to 2025. Among the profiled institutions, Instytut Farmakologii (yellow curve) exhibited the most significant growth, with a sharp increase in annual publications post-2016. Air Force Medical University, Instytut Farmakologii, Kaohsiung Medical University, and Nantong also demonstrated accelerated growth after 2010, while Seoul National University (SNU) (purple curve) maintained steady, moderate expansion throughout the period.

### 3.4 Author collaboration and publication patterns

Table 1 quantifies the influence of leading authors in NP research (2000–2025), featuring metrics including h-index, g-index, and total citation counts (TC). The h-index (a widely adopted bibliometric indicator) reflects consistent research productivity by measuring how many of an author’s papers have been cited at least h times[14]; the g-index, by contrast, prioritizes highly cited works to highlight impact in key research domains[13].For example, JI RR (rank 1) holds an h-index of 33 and a g-index of 37, alongside a TC of 11,671—these metrics underscore his sustained, high-impact contributions to the field since 2003. ZHANG Y (rank 2) demonstrates a stronger concentration of influential works: with an h-index of 28 and a g-index of 50 (the highest g-index among top authors), his output reflects a broader citation reach despite a later research start in 2007. Notably, WATKINS LR (rank 5) achieves a high TC of 6,052 with an h-index of 22, indicating that his work has garnered substantial attention in core research areas.

**Table 1:** Top 10 authors on astrocytes in neuropathic pain research (2000–2025)

| Rank | Author | h_index | g_index | m_index | TC | NP | PY_start | Articles | Articles Fractionalized |
| --- | --- | --- | --- | --- | --- | --- | --- | --- | --- |
| 1 | JI RR | 33 | 37 | 1.435 | 11671 | 37 | 2003 | 37 | 8.99 |
| 2 | ZHANG Y | 28 | 50 | 1.474 | 2528 | 57 | 2007 | 57 | 8.28 |
| 3 | WANG Y | 24 | 37 | 1.412 | 1512 | 54 | 2009 | 54 | 8.28 |
| 4 | MIKA J | 22 | 27 | 1.222 | 1974 | 27 | 2008 | 27 | 4.65 |
| 5 | WATKINS LR | 22 | 26 | 0.957 | 6052 | 26 | 2003 | 26 | 4.20 |
| 6 | ZHANG J | 21 | 34 | 1.050 | 1570 | 34 | 2006 | 34 | 5.66 |
| 7 | GAO Y | 20 | 32 | 1.111 | 3577 | 32 | 2008 | 32 | 5.83 |
| 8 | LI Y | 20 | 33 | 1.176 | 1169 | 46 | 2009 | 46 | 5.52 |
| 9 | WANG W | 19 | 26 | 1.056 | 919 | 26 | 2008 | 26 | 3.35 |
| 10 | GHELARDINI C | 18 | 24 | 1.385 | 1091 | 24 | 2013 | 24 | 3.25 |

**Abbreviation:**TC: Total Citations; NP: Number of Publications; PY\_start: Year of First Publication

Author collaboration networks (Figs 4A and 4B) reveal distinct collaborative clusters within the research community. In Fig 4A, multiple small-to-medium collaborative subgroups are observed (e.g., the cluster centered on Gaol,Yongjing; the European cluster led by Ghelardini,Carla), indicating relatively concentrated regional/team-based collaborations. Fig 4B further expands the scope, showing a denser and more interconnected collaboration network: core authors (e.g., Ji Rr, Watkins Lr, Mannelli Ld) act as bridges across multiple clusters, while subgroups (e.g., the Gao, Y J cluster, Zimmermann, M cluster) reflect specialized research teams with close internal collaborations. The annual publication and temporal citation trends of high-output authors (Fig 4C) show that core authors (e.g., ZHANG Y-, WANG Y-) maintained stable annual publication output from 2007 to 2023. For JI R R, two notable citation peaks (around 2015 and 2019) correspond to high-impact works in these periods; authors like ZHANG X- showed a gradual increase in annual output in recent years (post-2019), indicating growing research activity. The author productivity distribution (Fig 4D) follows a typical Pareto principle: the vast majority of authors (over 90%) published ≤ 5 documents, while only a small proportion of core authors contributed ≥ 10 documents. This skewed distribution (a rapidly declining curve in the low-document range) reflects the “long tail” feature of author productivity in this field—research output is concentrated among a small number of high-productivity scholars.

**Fig 4:**
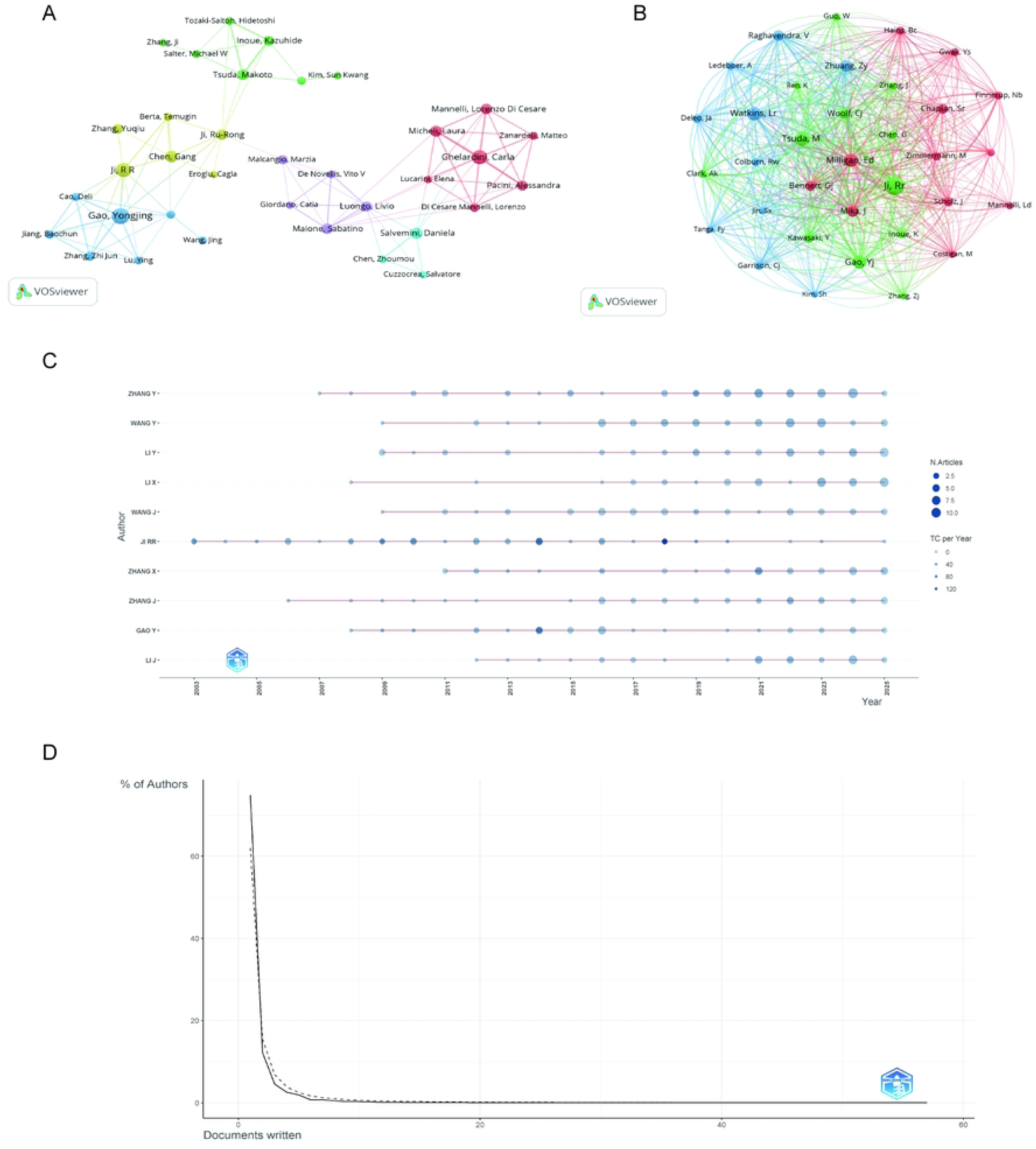
Visualization and analysis of author networks and co-cited authors in neuropathic pain. Co-authorship and co-cited authorship networks on astrocytes in the neuropathic field (2000–2025). (A) Co-authorship network: Nodes represent authors, sized by publication count. Colors indicate collaboration clusters. (B) Co-cited authorship network: Nodes, connected by collaborative publications, have edge thickness proportional to joint works. Clusters, marked by colors, reveal research groups. (C) Annual publication trends of the top 10 authors. (D) Lotka’s law fitting curve: Highlights the distribution of productivity and collaboration patterns among authors.

### 3.5 Journals and co-journals

The journal collaboration network (Figs 5A and 5B) reflects the pattern of cross-journal academic exchange pattern in this field. In Fig 5A, journals are clustered into three main groups (red, green, blue), with Pain as the core node (blue cluster) connecting multiple journals (e.g., *Eur J Pain*, *Exp Neurol*), indicating its central position in interdisciplinary exchanges. Table 2 further substantiates their scholarly influence. The leading status of these journals is demonstrated not only by their substantial publication output but also by their strong academic visibility and reputational standing. The majority are classified as Q1 journals in the JCR, underscoring their significant impact and broad recognition within the academic community. Fig 5B further expands the network: the green cluster (centered on the Journal of *Neuroscience* and *Pain*) dominates basic neuroscience and pain studies, with dense internal links. The red cluster (e.g., *Scientific Reports*, *Cells*) focuses on molecular/translational research, closely connecting with the green cluster. *Pain* acts as a core node bridging multiple subfields, reflecting the field’s integrated academic exchange.

**Fig 5:**
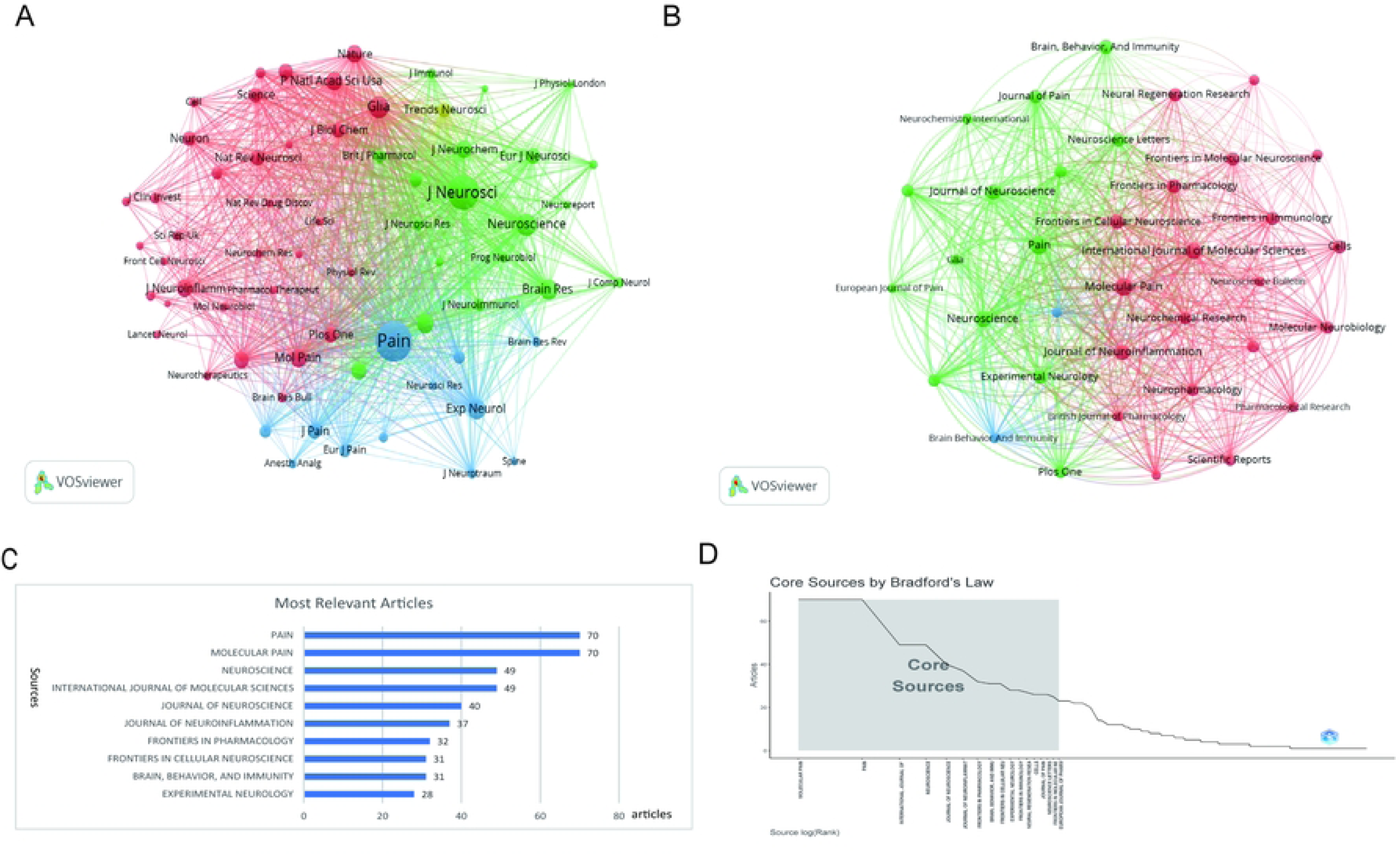
Visualization of journal association networks in neuropathic pain research. (A) Journal citation network: Nodes represent journals, connected by citations. Colors (green, blue, red) indicate thematic clusters of frequent inter-citation. (B) Journal co-citation network: Nodes, sized by co-citation frequency, are linked by co-citation ties. Colors denote clusters of commonly co-cited journals. (C) The X-axis shows article numbers, while the Y-axis lists the journals. (D) Bradford’s Law analysis: Identifies core journals based on publication distribution.

**Table 2:** Top 15 influential journals on astrocytes in neuropathic pain research (2000–2025)

| Rank | Source | h_index | g_index | m_index | TC | NP | PY_start | IF | JCR |
| --- | --- | --- | --- | --- | --- | --- | --- | --- | --- |
| 1 | PAIN | 44 | 70 | 1.76 | 6154 | 70 | 2001 | 5.5 | Q1 |
| 2 | JOURNAL OF<br>NEUROSCIENCE | 32 | 40 | 1.391 | 7089 | 40 | 2003 | 4 | Q1 |
| 3 | NEUROSCIENCE | 30 | 49 | 1.154 | 3312 | 49 | 2000 | 2.8 | Q1 |
| 4 | MOLECULAR PAIN | 28 | 43 | 1.556 | 2081 | 70 | 2008 | 2.8 | Q1 |
| 5 | JOURNAL OF<br>NEUROINFLAMMATION | 26 | 37 | 1.625 | 2510 | 37 | 2010 | 10 | Q1 |
| 6 | EXPERIMENTAL<br>NEUROLOGY | 24 | 28 | 1.143 | 2539 | 28 | 2005 | 4.2 | Q2 |
| 7 | BRAIN BEHAVIOR AND<br>IMMUNITY | 22 | 31 | 1.158 | 1968 | 31 | 2007 | 7.6 | Q1 |
| 8 | INTERNATIONAL JOURNAL<br>OF MOLECULAR SCIENCES | 22 | 35 | 2 | 1347 | 49 | 2015 | 4.9 | Q1 |
| 9 | JOURNAL OF PAIN | 19 | 26 | 0.792 | 1647 | 26 | 2002 | 4 | Q1 |
| 10 | FRONTIERS IN CELLULAR<br>NEUROSCIENCE | 18 | 31 | 1.385 | 1707 | 31 | 2013 | 4 | Q2 |
| 11 | JOURNAL OF<br>NEUROCHEMISTRY | 18 | 22 | 0.783 | 1690 | 22 | 2003 | 4 | Q2 |
| 12 | FRONTIERS IN<br>IMMUNOLOGY | 17 | 28 | 1.889 | 831 | 28 | 2017 | 5.9 | Q1 |
| 13 | EUROPEAN JOURNAL OF<br>PHARMACOLOGY | 16 | 23 | 0.889 | 1269 | 23 | 2008 | 4.7 | Q2 |
| 14 | FRONTIERS IN<br>PHARMACOLOGY | 16 | 28 | 1.778 | 795 | 32 | 2017 | 4.8 | Q1 |
| 15 | PLOS ONE | 16 | 20 | 0.941 | 1135 | 20 | 2009 | 2.6 | Q1 |

Abbreviation: IF: Impact Factor; JCR: Journal Citation Reports

The distribution of high-output journals (Fig 5C) shows that *Molecular Pain* and *Pain* are the most relevant sources (with 70 published documents each), followed by *International Journal of Molecular Sciences* and *Neuroscience* (with 49 documents). This indicates that top-tier journals in pain research and molecular neuroscience serve as the primary publication platforms for this field. Based on Bradford’s Law (Fig 5D), the journal distribution follows a segmented pattern: a small number of “Core Sources” (the shaded area) publish the majority of documents in this field, while the number of journals decreases sharply as publication volume declines. This result confirms that research output in this field is concentrated in a limited set of core journals, which play a leading role in academic dissemination.

### 3.6 Citation network and thematic evolution of neuropathic Pain research

The research on astrocytes in NP is influenced by prominent authors, high-impact journals, and highly cited studies. As shown in S1 Table, the analysis included 1,828 publications with 30,801 total citations (2000–2025), resulting in a mean age of 7.23 years and an average of 55.86 citations per article, indicating strong scholarly influence and sustained academic relevance.

The citation collaboration network (Fig 6A) illustrates the knowledge connection pattern of NP studies from 2000 to 2025. The ten most highly cited publications are summarized in Table 3. Core references (e.g., Ji RR (2019)[15], Donnelly CR (2020)[16], Finnerup NB (2021)[17]) are positioned at the center of the network, characterized by larger nodes and dense links, which indicate their high citation frequency and central status in academic discourse. Notably, Ji RR (with multiple nodes in 2013/2016/2018/2019) forms a key connection hub, reflecting the author’s sustained influence across different stages of research.

**Fig 6:**
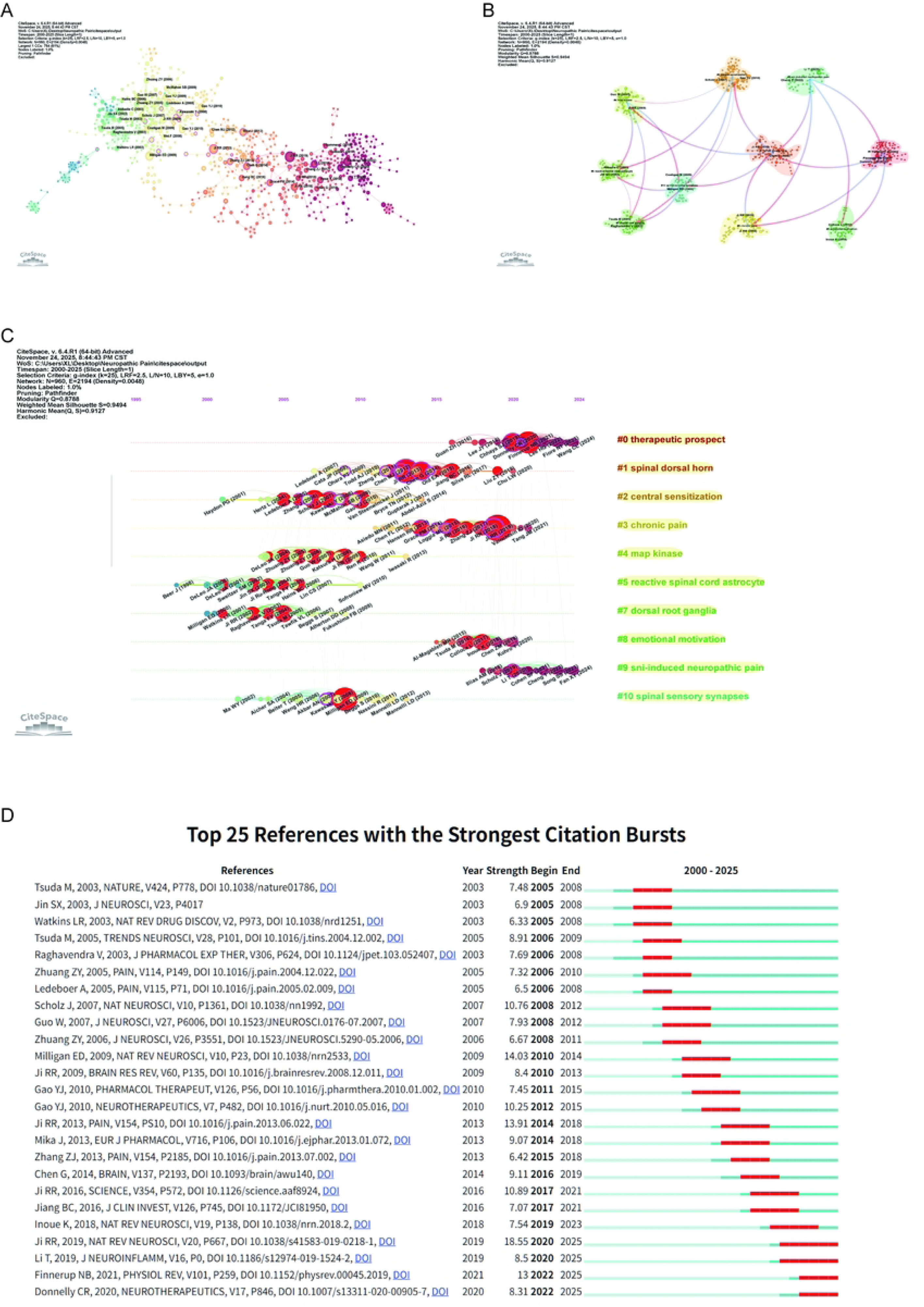
Network visualization of references in neuropathic pain. (A) Reference co-occurrence network based on CiteSpace, with nodes representing references, node size indicating frequency, and connections illustrating co-occurrence relationships. (B) Clustered co-citation network: Nodes, sized by citation frequency, are grouped by modularity with keyword labels for major clusters. Edge thickness reflects co-citation strength, while colors highlight thematic diversity. (C) Timeline clustering of publications. (D) Top 25 references with the strongest citation bursts in the research. The blue lines indicate the time intervals, while red lines mark the periods of burst activity for each reference, showing the initial and final years of the citation burst duration.

**Table 3:** Top 10 most cited publications based on bibliometrix analysis (2000–2025)

| RANK | First Author | Year | Journal | Paper | DOI | Total Citations | TC per Year | Normalized TC |
| --- | --- | --- | --- | --- | --- | --- | --- | --- |
| 1 | SOFRONIEW MV | 2010 | ACTA NEUROPATHOL | Astrocytes: biology and pathology | 10.1007/s00401-009-0619-8 | 4197 | 262.31 | 19.35 |
| 2 | RANSOHOFF RM | 2009 | ANNU REV IMMUNOL | Microglial Physiology: Unique Stimuli, Specialized Responses | 10.1146/annurev.immunol.021908.132528 | 1547 | 91.00 | 8.60 |
| 3 | SCHOLZ J | 2007 | NAT NEUROSCI | The neuropathic pain triad: neurons, immune cells and glia | 10.1038/nn1992 | 1534 | 80.74 | 8.15 |
| 4 | BURNSTOCK G | 2007 | PHYSIOL REV | Physiology and Pathophysiology of Purinergic Neurotransmission | 10.1152/physrev.00043.2006 | 1387 | 73.00 | 7.37 |
| 5 | MILLIGAN ED | 2009 | NAT REV NEUROSCI | Pathological and protective roles of glia in chronic pain | 10.1038/nrn2533 | 1231 | 72.41 | 6.84 |
| 6 | JI RR | 2018 | ANESTHESIOLOGY | Neuroinflammation and Central Sensitization in Chronic and Widespread Pain | 10.1097/ALN.0000000000002130 | 1103 | 137.88 | 19.92 |
| 7 | JI RR | 2014 | NAT REV DRUG DISCOV | Emerging targets in neuroinflammation-driven chronic pain | 10.1038/nrd4334 | 874 | 72.83 | 10.59 |
| 8 | JI RR | 2009 | BRAIN RES REV | MAP kinase and pain | 10.1016/j.brainresrev.2008.12.011 | 845 | 49.71 | 4.70 |
| 9 | JIN S | 2003 | J NEUROSCI | p38 Mitogen-Activated Protein Kinase Is Activated after a Spinal Nerve Ligation in Spinal Cord Microglia and Dorsal Root Ganglion Neurons and Contributes to the Generation of Neuropathic Pain | 10.1523/jneurosci.23-10-04017.2003 | 794 | 34.52 | 1.51 |
| 10 | GRACE PM | 2014 | NAT REV IMMUNOL | Pathological pain and the neuroimmune interface | 10.1038/nri3621 | 751 | 62.58 | 9.10 |

**Abbreviation:** DOI: Digital Object Identifier

The cluster dependency diagram (Fig 6B) identifies 10 distinct thematic clusters: #0 therapeutic prospect; #1 spinal dorsal horn & #7 dorsal root ganglia (anatomical basis of pain signal transmission); #2 central sensitization & #3 chronic pain (pathophysiological mechanisms of pain persistence); #4 map kinase (molecular signaling pathways); #5 reactive spinal cord astrocyte (glial cell role in pathogenesis); #8 emotional motivation (pain-psychological links); #9 SNI-induced neuropathic pain (experimental models); #10 spinal sensory synapses (synaptic transmission). Cross-cluster links (e.g., central sensitization-spinal dorsal horn) reflect the interdisciplinary integration of mechanisms and anatomical targets.

The cluster timeline (Fig 6C) illustrates the temporal thematic evolution: 2000–2010 (early stage), clusters (#7, #10) established anatomical/molecular foundations; 2010–2020 (mid-stage) saw the rise of functional mechanism clusters (#2, #4); 2020–2025 (recent stage) shifted to translational research and model optimization (#0, #9). Ji RR contributed to multiple clusters over time, underscoring continuous contributions to key fronts.

Fig 6D ranks the top 25 references by citation burst strength. Milligan ED (2009)[18] demonstrated the strongest burst (strength=14.03, 2010–2014), laying a foundation for early 2010s research. High-impact references also include Ji RR (2019)[15], (strength=18.55, 2020–2025) and Scholz J (2007)[19], (strength=10.76, 2008–2012). Early bursts (e.g., Tsuda M (2003)[20], and Zhuang ZY (2005)[21]) focused on basic mechanisms, while mid-to-late bursts (e.g., Ji RR (2016)[22], and Finnerup NB (2021)[17] leaned toward translation and clinical research. These bursts trace the field’s evolution from basic mechanisms to translational applications, with core scholars like Ji RR driving high-impact fronts.

### 3.7 Keywords analysis

Keyword analysis identifies core themes and evolutionary trends in NP research (2000–2025) via VOSviewer and CiteSpace. A total of 11,601 Keywords Plus and 3,576 author keywords were analyzed, comprehensively characterizing research foci (S1 Table).

As shown in Figs 7A and 7C, “neuropathic pain” (term frequency=1393, total link strength=1427) is the dominant central theme, closely associated with “astrocytes” (frequency=1718, link strength=1174) and “microglia” (frequency=906, link strength=952). Other high-frequency terms include “spinal cord” (490) and “neuroinflammation” (426). Fig 7B reveals dynamic shifts: pre-2016 focus on model-based terms (e.g., “chronic constriction injury”) shifted post-2018 to “oxidative stress” and “diabetic neuropathic pain,” expanding into comorbid and mechanistic dimensions. Fig 7D (trend topic plot, 2004–2024) illustrates temporal thematic evolution: 2004–2012 (early phase) centered on basic inflammatory mediators (e.g., tumor necrosis factor-alpha, interleukin-1) with low frequencies (<200); 2012–2018 (mid phase) saw rapid growth of core themes (e.g., neuropathic pain, astrocytes, frequency>400); 2018–2024 (late phase) witnessed surging frontier topics (e.g., electroacupuncture, reactive oxygen species, frequency>600), reflecting an interest in non-pharmacological therapies and novel mechanisms.

**Fig 7:**
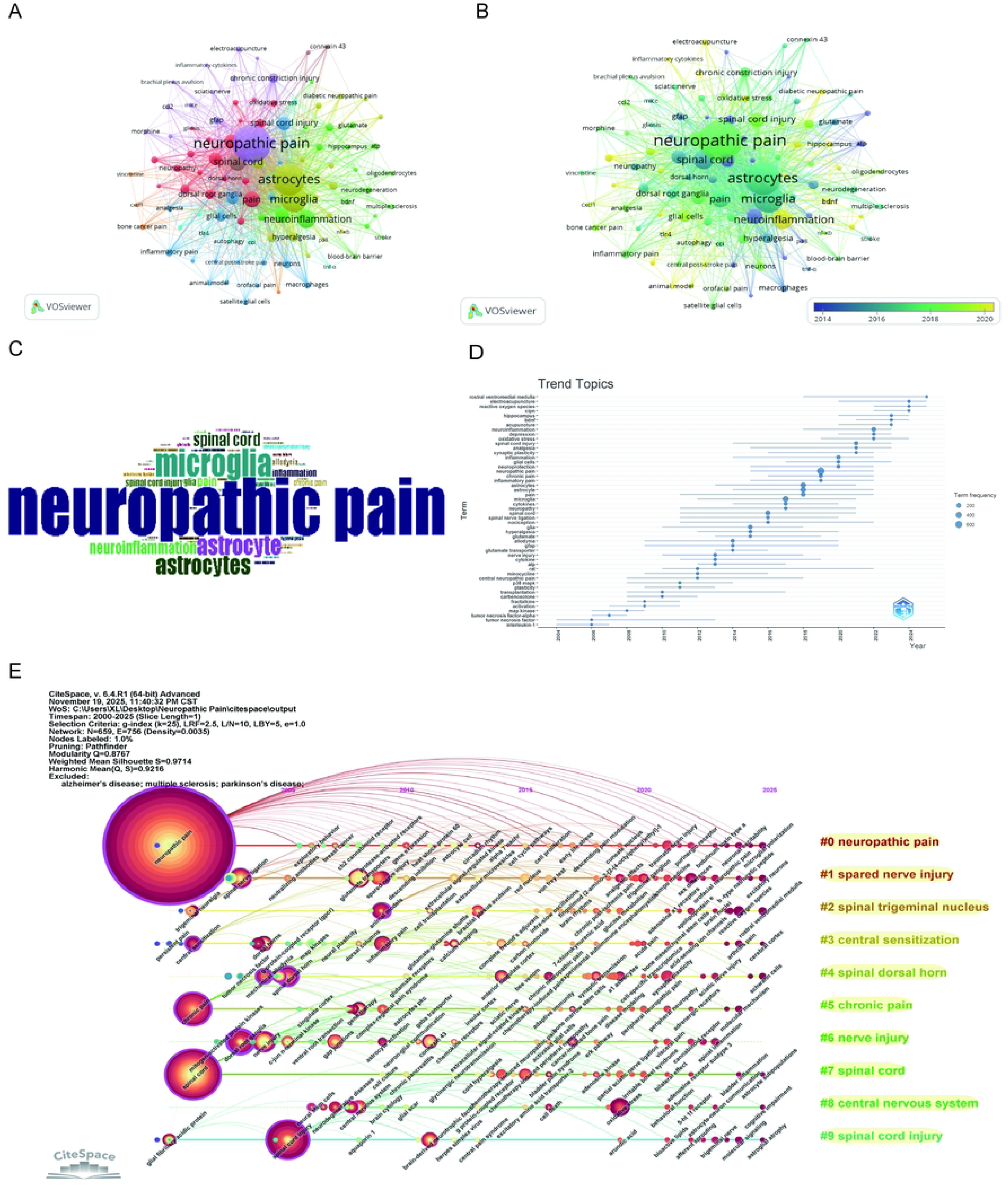
Visualization of keywords in neuropathic pain research. (A) Co-occurrence Clustering of Keywords and (B) Average Normalized Citations in Author-Keyword Analysis. Each circle and its associated label represent a keyword unit, with different colors indicating distinct clusters. (C) Author Keywords Word Cloud: The size of each word reflects its frequency. (D) Keywords Timeline: Nodes, sized by frequency within each cluster, illustrate the temporal evolution of research topics. (E) Co-citation Timeline: Clusters are shown as horizontal lines, with node size reflecting co-citation frequency, links indicating relationships, and node positions marking the first co-citation year.

Using CiteSpace for keyword burst analysis (Fig 8), we can delineate temporal shifts in research hotspots and cutting-edge directions in NP studies (2000–2025), while identifying emerging trends. Based on burst timing and intensity, the top 25 keywords with the strongest citation bursts can be categorized into three phases, reflecting distinct stages of academic evolution.In the early phase (2000–2010), research focused on foundational mechanisms and anatomical correlates of pain. Keywords like glial fibrillary acidic protein (burst strength=2.23, 2000–2011) and dorsal root ganglia (burst strength=2.59, 2003–2007) exhibited significant bursts, highlighting early attention to glial cell markers and peripheral pain-signaling structures. Nerve injury (burst strength=5.73, 2004–2013) emerged as a core theme, underscoring the field’s initial focus on injury-induced pain models and basic pathophysiology.The middle phase (2011–2020) saw a shift toward mechanistic and therapeutic exploration. Glutamate transporters (burst strength=5.04, 2010–2016) and chronic constriction injury (burst strength=3.6, 2014–2019) gained traction, reflecting deepened interest in synaptic neurotransmission and preclinical pain models. Satellite glial cells (burst strength=2.68, 2017–2020) and oxidative stress (burst strength=3.91, 2019–2025) also exhibited late-phase bursts during this period, signaling a growing focus on glial-mediated inflammation and redox-related pain mechanisms.By the late phase (2021–2025), the field shifted towards translational and specialized subdomains. Synaptic plasticity (burst strength=3.27, 2021–2022) and sex differences (burst strength=3.17, 2022–2025) emerged as new areas of interest, emphasizing the significance of circuit-level pain adaptation and personalized pain responses. Notably, spinal cord injury (burst strength=5.39, 2023–2025) exhibited the strongest late-phase burst, reflecting a renewed focus on comorbid pain in neurological trauma—aligning with clinical demands for integrated injury-pain management.

**Fig 8:**
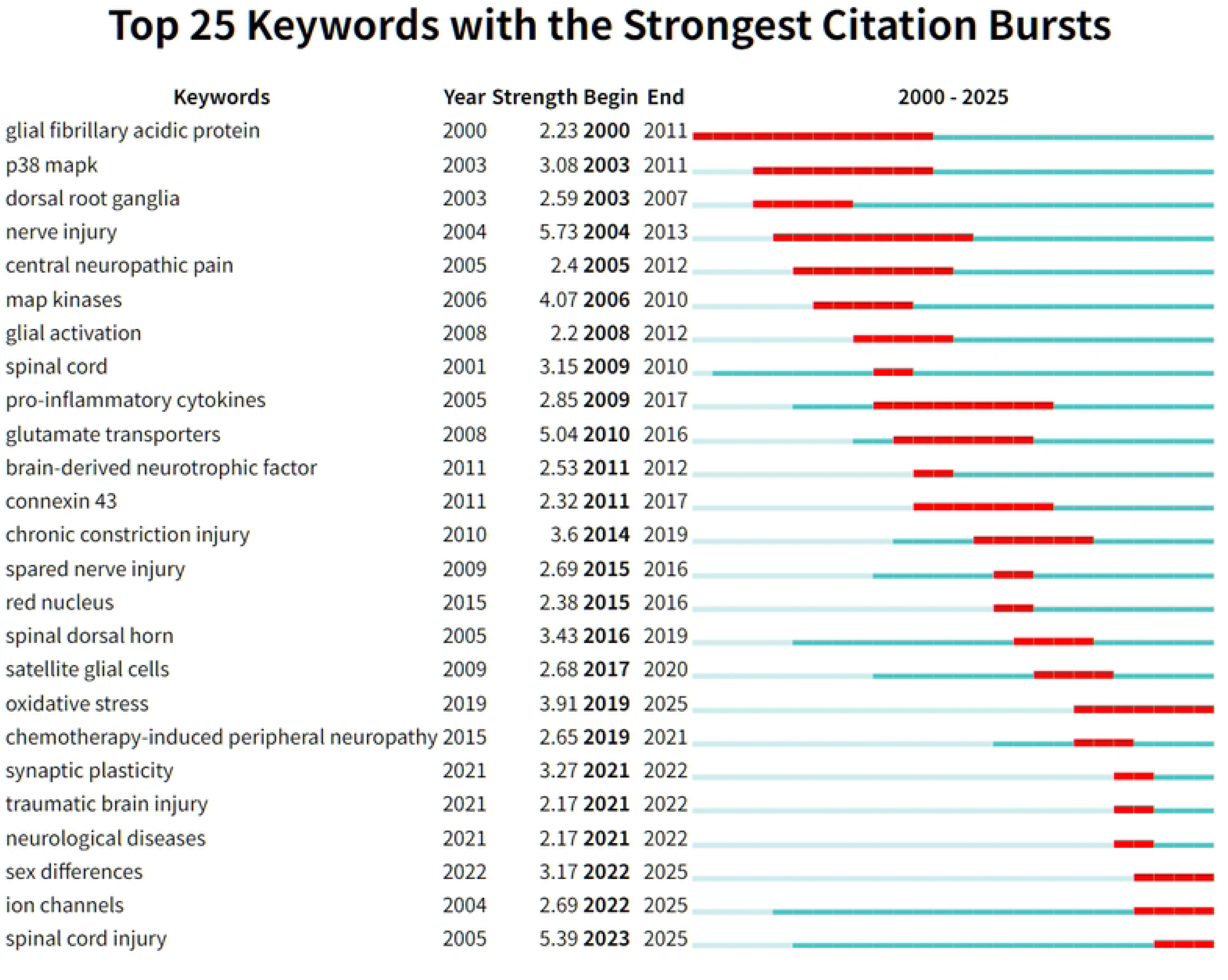
Top 25 keywords with the strongest citation bursts in neuropathic pain research. The burst timeline (right-hand side) displays the periods during which these bursts occurred, with red segments indicating the time intervals when each reference received heightened attention. The burst detection process highlights key works that had a significant impact in a short time, suggesting pivotal contributions to the evolving research landscape in neuropathic pain.

## 4. Discussion

This study employs bibliometric analysis to explore the scientific landscape of NP and astrocytes, addressing the lack of comprehensive insights into research trends and therapeutic targets. The analysis reveals a significant increase in publications and citations, highlighting the growing interest in the roles of astrocytes in NP pathogenesis. Key findings include the identification of influential authors, collaborative networks, and thematic evolution in research, with a focus on potential mechanisms and therapeutic strategies. The study underscores the need for targeted therapies that address astrocytic dysfunction, providing a roadmap for future investigations in this critical area of pain research.

### 4.1 Reactive astrocytes

Astrocytes, a type of glial cell in the central nervous system, play a significant role in the pathophysiology of NP. These cells express glial fibrillary acidic protein (GFAP), a marker of astrocyte activation, which is often elevated in conditions associated with NP.

Reactive astrocytes, identified by elevated GFAP levels, exhibit different states of reactivity that are intricately linked to the development and maintenance of NP. For instance, in a rat model of NP induced by spinal nerve ligation, increased GFAP expression was observed alongside the development of mechanical allodynia, a common symptom of NP, suggesting that activated satellite glial cells (SGCs) marked by GFAP contribute to the maintenance of NP[23]. Similarly, in Fabry-Anderson disease, NP is associated with severe neuroinflammation in the dorsal root ganglia, characterized by the overexpression of GFAP, indicating a significant role of glial activation in the pathophysiology of pain[24]. For example, in the context of syringomyelia and NP in Cavalier King Charles Spaniels, reactive astrocytes in the dorsal horn exhibit varied states of reactivity, suggesting a complex role in NP pathogenesis[5].The CXCL13/CXCR5 chemokine pathway is another mechanism through which reactive astrocytes contribute to NP by promoting central sensitization and pain hypersensitivity; inhibiting this pathway significantly reduces pain in preclinical models[25]. Additionally, the expression of iNOS in reactive astrocytes has been linked to the maintenance of NP, as evidenced by increased iNOS levels in the spinal cord of rats with spinal cord injury[26]. Reactive astrocytes also undergo subpopulation changes in response to nerve injury, which are associated with the development of NP, highlighting their role in nociceptive signaling[27]. These findings underscore the involvement of astrocytes and GFAP in the mechanisms underlying NP, emphasizing the potential of targeting astrocytic activation as a therapeutic strategy for alleviating NP symptoms.

### 4.2 Neuroinflammatory signaling

The activation of astrocytes is not only a response to injury but also a driver of neuroinflammation, as observed in conditions such as Parkinson’s disease and Alzheimer’s disease, where astrocytes mediate inflammatory processes that exacerbate disease progression[28]. Following various types of neural injury, such as optic nerve crush or traumatic brain injury, astrocytes become activated and contribute to the neuroinflammatory response by upregulating genes related to inflammation and releasing pro-inflammatory cytokines[29, 30]. In chronic pain conditions, reactive astrocytes transition from a neuroprotective to a neurotoxic phenotype, further promoting neuroinflammation and contributing to the persistence of pain[31].

Neuroinflammation, characterized by the release of pro-inflammatory cytokines and oxidative stress, is a key factor in the pathophysiology of NP. Pro-inflammatory cytokines such as TNF-α, IL-1β, and IL-6 are elevated in various models of NP, contributing to the sensitization of pain pathways and the persistence of pain symptoms[32, 33]. Oxidative stress further exacerbates neuroinflammation by disrupting cellular homeostasis and promoting the activation of inflammatory pathways, which are implicated in the development of NP[34, 35]. The interplay between oxidative stress and pro-inflammatory cytokines not only drives the progression of NP but also highlights potential therapeutic targets. For instance, interventions that reduce oxidative stress or modulate cytokine production have shown promise in alleviating NP symptoms[36, 37].

Overall, the interaction between neuroinflammation, pro-inflammatory cytokines, and oxidative stress underscores the complexity of NP and the need for multifaceted therapeutic approaches.

### 4.3 Glutamate homeostasis and synaptic plasticity

The interaction between NP and astrocytes is significant in the context of pain modulation. Astrocytes play a crucial role in maintaining synaptic plasticity and glutamate homeostasis, which are essential for normal neuronal function. In the case of NP, astrocytes are involved in modulating pain through the NDRG2/GLT-1 pathway, which reduces synaptic glutamate levels and normalizes synaptic plasticity, thereby alleviating pain symptoms[38]. This highlights the importance of astrocytes in regulating glutamate transporters, which are critical for maintaining synaptic balance and preventing excitotoxicity.

Glutamate transporters, particularly EAAT2, are essential for regulating extracellular glutamate levels, which in turn influence synaptic plasticity. In NP, the regulation of glutamate transporters is crucial for modulating synaptic changes that contribute to pain persistence. For instance, treatments that enhance glutamate reuptake, such as ceftriaxone, have been shown to increase pain thresholds by upregulating glutamate transporters like GLT-1, thereby reducing excitatory neurotransmission and alleviating chronic pain[39]. Additionally, synaptic plasticity is affected by the modulation of glutamate transporters, as demonstrated by the normalization of synaptic function through the astrocytic NDRG2/GLT-1 pathway, which underscores the interconnectedness of these elements in the context of NP[38].

### 4.4 Emerging frontiers: comorbidities, and personalized pain

Astrocytes are involved in neuroinflammation and synaptic dysfunction, which are key mechanisms in the development and maintenance of NP. However, comorbidities and sex differences significantly influence the experience and management of NP. Comorbid conditions such as depression, anxiety, and cardiovascular disease are frequently associated with NP, complicating its prognosis and treatment[40, 41]. For example, depression and insomnia serve as significant mediators in the relationship between problematic internet use and NP, indicating the need for integrated treatment strategies[42]. Sex differences also play a crucial role, with studies showing that female patients often experience a higher prevalence of NP and may respond differently to treatments compared to males[43, 44]. For instance, cannabinoid-based therapies have been found to be more effective in female patients, underscoring the necessity for sex-specific approaches in analgesic development[44]. These insights emphasize the importance of considering both comorbidities and sex differences in the clinical management of NP.

### 4.5 Limitations

Despite offering a thorough examination of publication patterns and collaborative structures within the field, this study has several acknowledged limitations. First, the analysis is largely based on records obtained from WoSCC and Scopus, which may not fully capture all pertinent literature in this domain. Second, the scope of this study was restricted to English-language literature, potentially leading to the underrepresentation of significant research contributions published in other languages and regions. Additionally, a noticeable geographical imbalance was observed in the distribution of research output, with the United States and China dominating the field. This imbalance suggests that the findings may not fully reflect the global landscape of research activity and investment. To address these limitations, future studies should broaden the range of data sources by incorporating additional bibliographic databases and including multilingual and regionally diverse literature, thereby providing a more comprehensive and balanced assessment of global scholarly influence in this field.

## 5. Conclusion

Our study underscores the central role of astrocytes in NP and offers a comprehensive framework to guide future mechanistic research and the development of astrocyte-targeted therapeutic strategies. The future direction of NP research is shifting towards a deeper understanding of the complex molecular and cellular mechanisms underlying the condition, with a strong emphasis on translating these insights into more effective and personalized treatments.

## Acknowledgments

The authors would like to thank the editors and the anonymous reviewers for their valuable comments and suggestions, which helped improve the quality of the paper.

## Statements and declarations

### Author contributions

Li Xue (Data curation; Formal analysis; Methodology; Software; Writing – original draft; Writing – review & editing); Haixia Fan (Writing – review & editing); Hong Yuan (Software; Supervision; Visualization); Jiao Yang (Software; Supervision; Visualization); Qiurui Yuan (Software; Supervision; Visualization).

### Data Availability

The data supporting the findings of this study are available upon request from the corresponding author.

## Abbreviations

EAAT2: Excitatory Amino Acid Transporter 2
GFAP: Glial Fibrillary Acidic Protein
GLT-1: Glutamate Transporter-1
IL-1β: Interleukin-1 Beta
IL-6: Interleukin-6
iNOS: Inducible Nitric Oxide Synthase
MeSH: Medical Subject Headings
NC: Number of Citations
NP: Neuropathic Pain
NDRG2: N-Myc Downstream Regulated Gene 2
SGCs: Satellite Glial Cells
TC: Total Citations
TNF-α: Tumor Necrosis Factor-Alpha
WoSCC: Web of Science Core Collection.

## Supporting information

**S1 Table.** Bibliometric overview of astrocytes in neuropathic pain (2000–2025)

**S2 Table.** Most relevant countries by corresponding author contributions (2000–2025)

**S3 Table.** TOP 10 affiliations and article counts on neuropathic Pain (2000–2025)

**S4 Table.** Data search expressions

